# Feasibility and diagnostic accuracy of saliva-based SARS-CoV-2 screening in educational settings and children aged < 12 years

**DOI:** 10.1101/2021.04.17.21255651

**Authors:** Martin Hoch, Sebastian Vogel, Ute Eberle, Laura Kolberg, Valerie Gruenthaler, Volker Fingerle, Nikolaus Ackermann, Andreas Sing, Bernhard Liebl, Johannes Huebner, Simone Kuttiadan, Anita Rack-Hoch, Melanie Meyer-Buehn, Tilmann Schober, Ulrich von Both

**Author notes:** **Corresponding author:** Ulrich von Both, M.D., FRCPCH, Hauner Children’s Hospital, LMU Ludwig-Maximilians-University, Lindwurmstr. 4, 80337, Munich, Germany. These authors contributed equally to this manuscript (Martin Hoch, Sebastian Vogel, Ute Eberle).

## Abstract

Children have been disproportionately affected during the COVID-19 pandemic. We aimed to assess a saliva-based algorithm for SARS-CoV-2 testing to be used in schools and childcare institutions under pandemic conditions. A weekly SARS-CoV-2 sentinel study in primary schools, kindergartens and childcare facilities was conducted over a 12-week-period. In a sub-study covering 7 weeks, 1895 paired oropharyngeal and saliva samples were processed for SARS-CoV-2 rRT-PCR testing in both asymptomatic children (n=1243) and staff (n=652). Forty-nine additional concurrent swab and saliva samples were collected from SARS-CoV-2 infected patients (patient cohort). The Salivette® system was used for saliva collection and assessed for feasibility and diagnostic performance. For children a mean of 1.18 ml saliva could be obtained. Based on results from both cohorts, the Salivette® testing algorithm demonstrated specificity of 100% (95% CI 99.7 - 100) and sensitivity of 94.9% (95% CI 81.4 - 99.1) with oropharyngeal swabs as reference. Agreement between sampling systems was 100% for moderate to high viral load situations (defined as Ct-values < 33 from oropharyngeal swabs). Comparative analysis of Ct-values derived from saliva vs. oropharyngeal swabs demonstrated a significant difference (mean 4.23; 95% CI 2.48–6.00). In conclusion, the Salivette® system proved to be an easy-to-use, safe and feasible saliva collection method and a more pleasant alternative to oropharyngeal swabs for SARS-CoV-2 testing in children aged 3 years and above.

## Introduction

Children, in particular the group of <12-year-olds, are known to be at reduced risk for suffering from COVID-19. However, they have been substantially affected by closures of schools, kindergartens and childcare facilities in the ongoing pandemic [1,2]. Among many other reports, a recent commentary by Nagakumar and colleagues highlights the urgent need to develop strategies to safely operate schools while avoiding their closures [3]. Hence, scientists and public health leaders alike have been exploring options for coronavirus testing approaches in educational settings. Accumulating evidence points towards a rather low and stable transmission risk in educational institutions despite rising incidence rates in the population, as long as preventative hygiene measures and regular testing strategies are in place [4]. The ideal system to allow for large-scale test operations would be child-friendly, safe to perform, and ideally allow for self-sampling at home or at the appropriate child-care institution without the help of a medical professional. Various groups have explored different sampling methods from a range of clinical specimens while naso-/ oropharyngeal swabs are considered the gold standard [5–8]. Since no discernable differences in viral loads or detection rates had been demonstrated between naso- and oropharyngeal swabs, oropharyngeal swabs have been considered the primary choice for SARS-CoV-2 testing in children to minimize injuries on the nasopharyngeal route [6,9]. Numerous reports have described saliva sampling in adults as a reliable non-invasive method for SARS-CoV-2 testing with a sensitivity of > 80% and a specificity of >95% compared to naso-/oropharyngeal swabs [10–15]. Only very few reports have addressed saliva sampling in pediatric cohorts [16]. Of note, two studies comparing naso-/oropharyngeal swabs and saliva samples in symptomatic children found an overall saliva sensitivity of 85.2% (up to 95.2% in patients with high viral load (≥10^4^ copies/ml in nasopharyngalswabs) and 87.7%, respectively [17,18]. The Salivette® system has been proposed as a device for collecting saliva for SARS-CoV-2 testing in adults [19–22]. However, feasibility and diagnostic performance of this system in children and for routine testing in educational settings have not been assessed. Hence, the aim of our study was to establish and assess a practical, safe and easy-to-use system for saliva collection in educational settings using the Salivette® and subsequent rRT-PCR testing for SARS-CoV-2 for adult staff and children aged 3 years and above.

## Methods

### 2.1 Study setting and cohorts

Between June and November 2020, we conducted a weekly SARS-CoV-2 sentinel study in primary schools, kindergartens and childcare facilities in Munich following approval by the Ethics Committee of the Ludwig-Maximilians University Munich [4]. In a sub-study covering a 7-week period a total of 1895 paired oropharyngeal and saliva samples were obtained from asymptomatic children and staff attending participating educational institutions. Written informed consent was obtained from all individuals participating in this study. The Salivette® system was used for saliva sampling (SARSTEDT AG & Co KG, Nuembrecht, Germany; product number 51.1534). Over the 7-week period, we collected concurrent saliva and oropharyngeal swab sample pairs from children (n = 1243) and adult staff (n = 652) – sentinel cohort (SC, n = 1895). In parallel, 49 individuals, both adults and children known to be infected with SARS-CoV-2, were recruited and consented for prospectively paired Salivette® and oropharyngeal swab sampling - patient cohort (PC, n = 49). PC participants were either recruited in the hospital inpatient setting or in collaboration with public health services by visiting quarantined individuals at home. To identify eligible individuals, the study team was notified of a positive PCR test for SARS-CoV-2 obtained during routine clinical sampling.

### 2.2 Salivette® sampling and laboratory processing

Supervised saliva sampling and swabbing was performed by medically trained study personnel in specifically assigned rooms on site at the participating educational institu-tions. Samples were obtained after a minimum of 30 minutes since the last food and drink intake. Participants were asked to leave the Salivette®’s absorbent cotton pad in their mouth for a minimum of 2 minutes; subsequently, each individual replaced the pad into the Salivette® collection tube and closed it with the topper. Concurrent oropharyngeal swabs were taken immediately after saliva sampling. Following collection, samples were immediately transferred into the study laboratory. Salivette® tubes were centrifuged for 5 minutes at 1600g at room temperature to harvest saliva, following a protocol previously established in our children’s hospital’s routine diagnostic laboratory. A subset of samples was measured for saliva volume obtained by individual pipetting. All saliva specimens and swabs were processed using the ampliCube Coronavirus SARS-CoV-2 (Mikrogen, Germany) on a Bio-Rad CFX96 Touch rRT-PCR Detection System (Bio-Rad, Germany). Single gene results were retested with Xpert Xpress SARS-CoV-2 (Cepheid, USA). For methodological comparison between swab and saliva sampling we referred to semi-quantitative cycle threshold (Ct) values of corresponding SARS-CoV-2 gene locus.

### 2.3 Statistical analysis

Statistical analysis was done using R-studio software, version 4.0.2.3 for chi-square test, Wilcoxon signed rank test with continuity correction and Bland-Altman graphical analysis [23].

## Results

### 3.1 Feasibility of Salivette® sampling and pre-analytic aspects

We were able to fully standardize saliva sampling by using the Salivette®. Risk for viral spreading was found to be negligible due to the closed collection system compared to more open systems (spitting, gargling) potentially producing aerosols. Furthermore, the tubes require minimal storage space and are compatible with standard centrifuges making large-scale laboratory processing very feasible. A subset of 875 individual samples (574 children, 301 staff) were subjugated to accurate measurements of saliva volume to explicitly address pre-analytic aspects. We found that for children across all age groups a mean of 1,18 ml saliva could be obtained. For staff of the participating institutions a mean of 1.34 ml saliva could be collected (Table 1).

**Table 1:** Maximum, mean and minimum amount of saliva collected using the Salivette® system in children and staff (n=875): Volume (ml).

| Volume [ml] | 3-4 years<br>(n=145) | 5-6 years<br>(n=167) | 7-8 years<br>(n=170) | 9-11 years<br>(n=92) | All children<br>(n=574) | Staff<br>(n=301) |
| --- | --- | --- | --- | --- | --- | --- |
| Maximum | 2.50 | 2.50 | 3.00 | 2.50 |  | 2.75 |
| Mean | 1.04 | 1.13 | 1.21 | 1.33 | 1.18 | 1.34 |
| Minimum | 0.20 | 0.20 | 0.20 | 0.25 |  | 0.20 |

### 3.2 Salivette® diagnostic performance

Of 1895 prospectively paired oropharyngeal swab and Salivette® samples collected in the sentinel cohort (SC), 1893 showed a negative and 2 samples yielded a positive result. Thus, as expected, the SC proved to be a low incidence cohort with only two positive pairs of samples detected in week 12 [4]. It therefore only allowed for accurate assessment of specificity of the Salivette® method in relation to oropharyngeal swabs. As a consequence, we chose to establish an additional cohort (patient cohort, PC, n= 49) characterized by a high pre-test probability to evaluate sensitivity of the Salivette® testing method. Median age of this group was 46 years (range 3 to 87 years, male/female ratio 1.3) and assessment of saliva volume per Salivette® showed a mean of 1.73 ml (range: 0.75 – 2.75 ml). A total of eight individuals in the PC tested negative in both saliva and oropharyngeal swab samples. Thirty-seven individuals showed a positive test result from both sampling materials. Finally, two individuals demonstrated a discordant negative/positive and two additional individuals showed a discordant positive/negative result for Salivette® and oropharyngeal swab samples, respectively. For negative saliva samples Ct-values from corresponding oropharyngeal swab samples were 33.17 and 33.72, while for negative oropharyngeal swab samples Ct-values from corresponding positive saliva samples read 37.49 and 37.68, respectively. Based on combined results from both cohorts, the Salivette® testing method could be assigned a specificity of 100 % (95% CI 99.7 - 100) and a sensitivity (percentage of positive agreement) of 94.9 % (95% CI 81.4 - 99.1) in relation to oropharyngeal swabs (Table 2 a and b).

**Table 2a & b:**
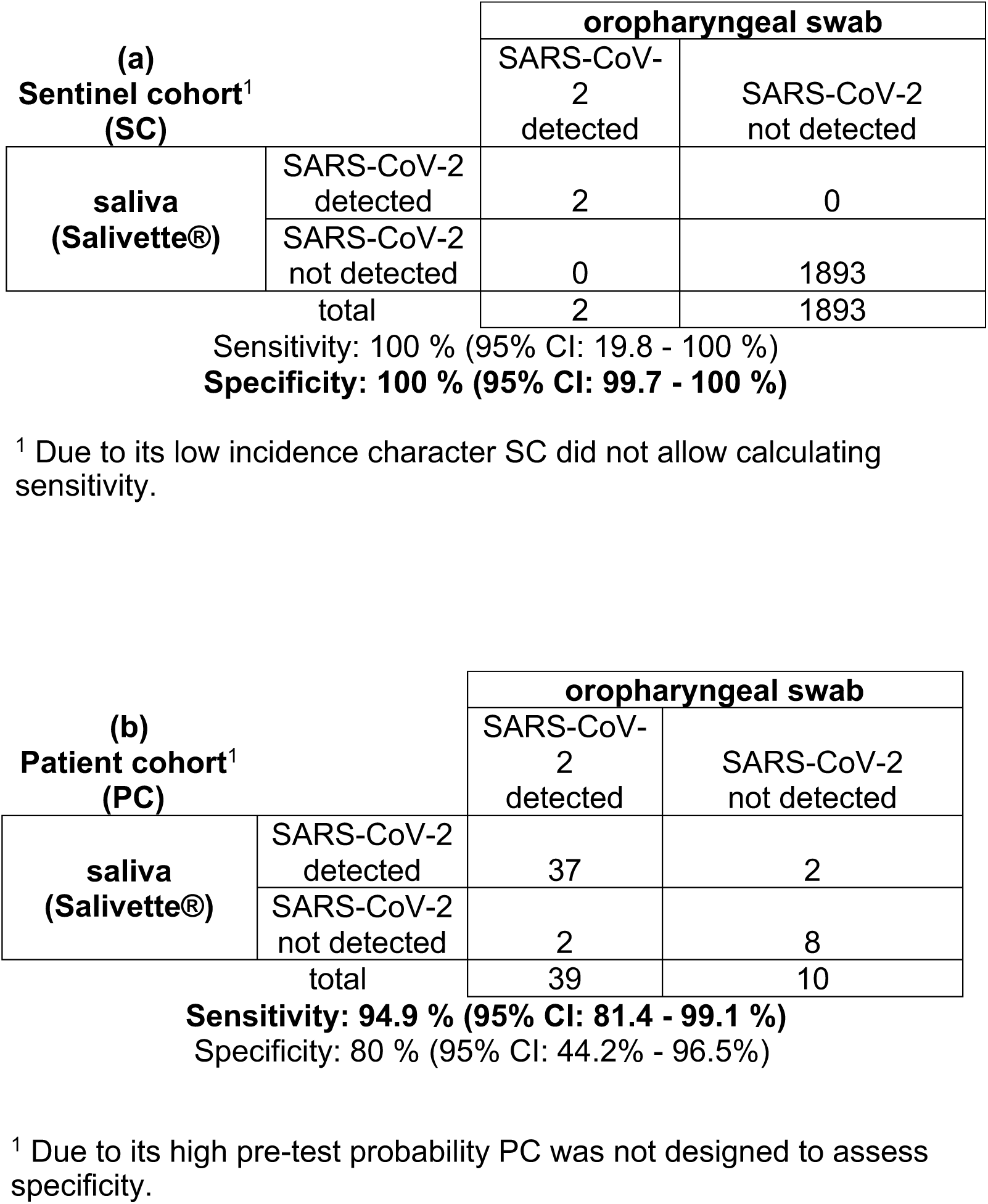

### 3.3 Ct-value comparison

To describe the effect of saliva sampling on Ct-value in comparison to oropharyngeal swabs, we assessed all Ct-values of individual sample pairs. Figures 2 and 3 visualize patient-matched saliva and swab SARS-CoV-2 rRT-PCR Ct-values for respective 39 cor-responding sample pairs (2 from SC and 37 from PC). Wilcoxon signed rank test with continuity correction showed a significant difference between Ct-value measurements derived from saliva vs. oropharyngeal swabs (p-value = 0.032). In addition, Bland-Altmann graphical comparison showed agreement between the two sampling methods with saliva-derived Ct-values being systematically higher than Ct-values derived from oropharyngeal swabs: Mean difference 4.23 (95% CI 2.48–6.00), upper limit of agreement 14.85 (95% CI 17.87 – 11.82) and lower limit of agreement -6.38 (95% CI -9.41 – -3.35) [23]. Ct-value data for all 39 sample pairs, including age distribution of individuals tested, are listed in the supplementary material (Table S1). In addition, we separately analyzed results of corresponding sample pairs for individuals with a moderate to high viral load (defined as Ct-values < 33 from oropharyngeal swab samples) and were able to demonstrate that positive percent agreement between the two sampling systems was 100%.

**Figure 1.**
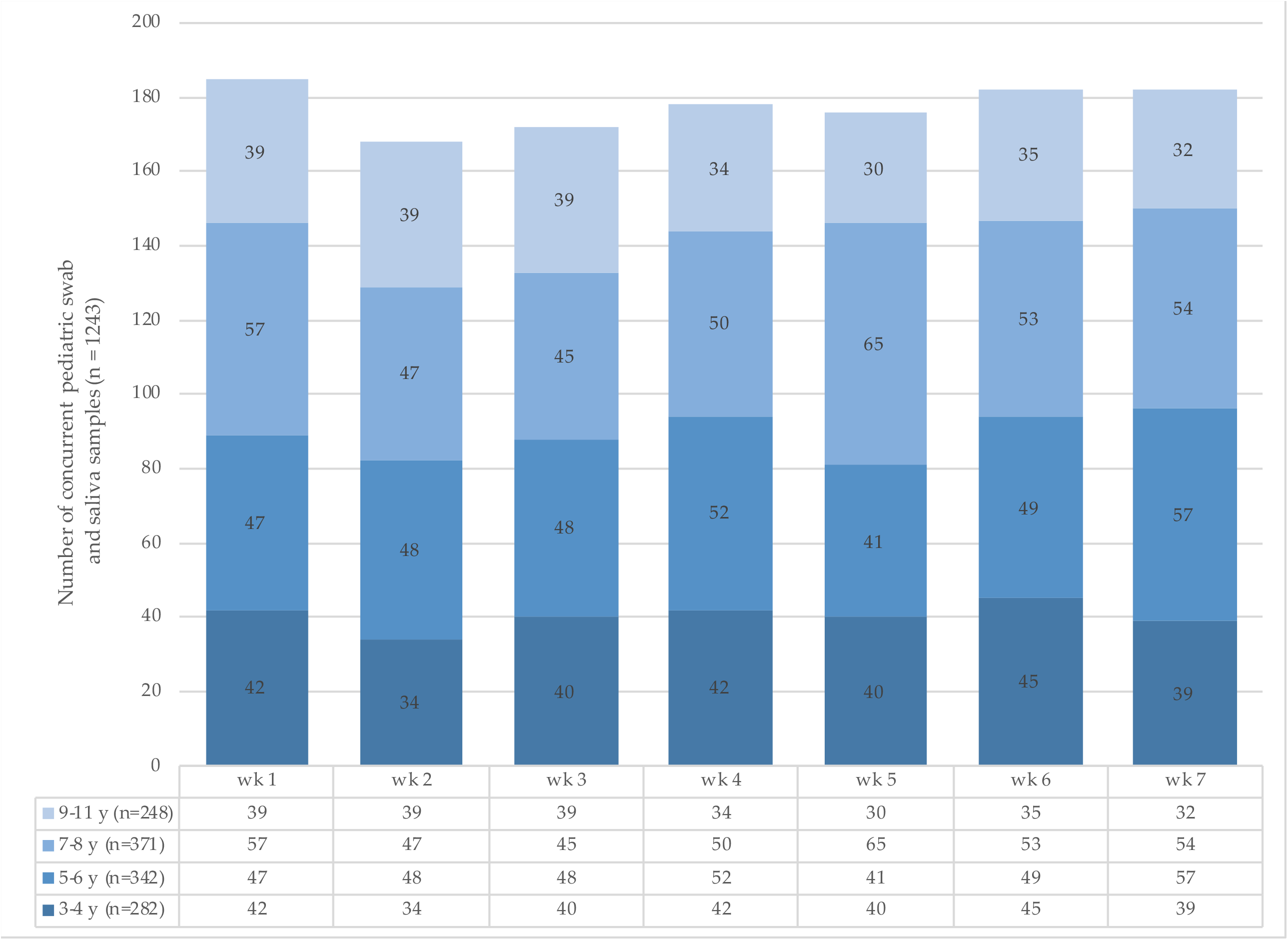
Pediatric sample pairs (oropharyngeal swab and Salivette®) collected for SARS-CoV-2 rRT-PCR testing per study week. Colored bars illustrate stratification for individual age groups starting from the age of 3 years. y = years.

**Figure 2.**
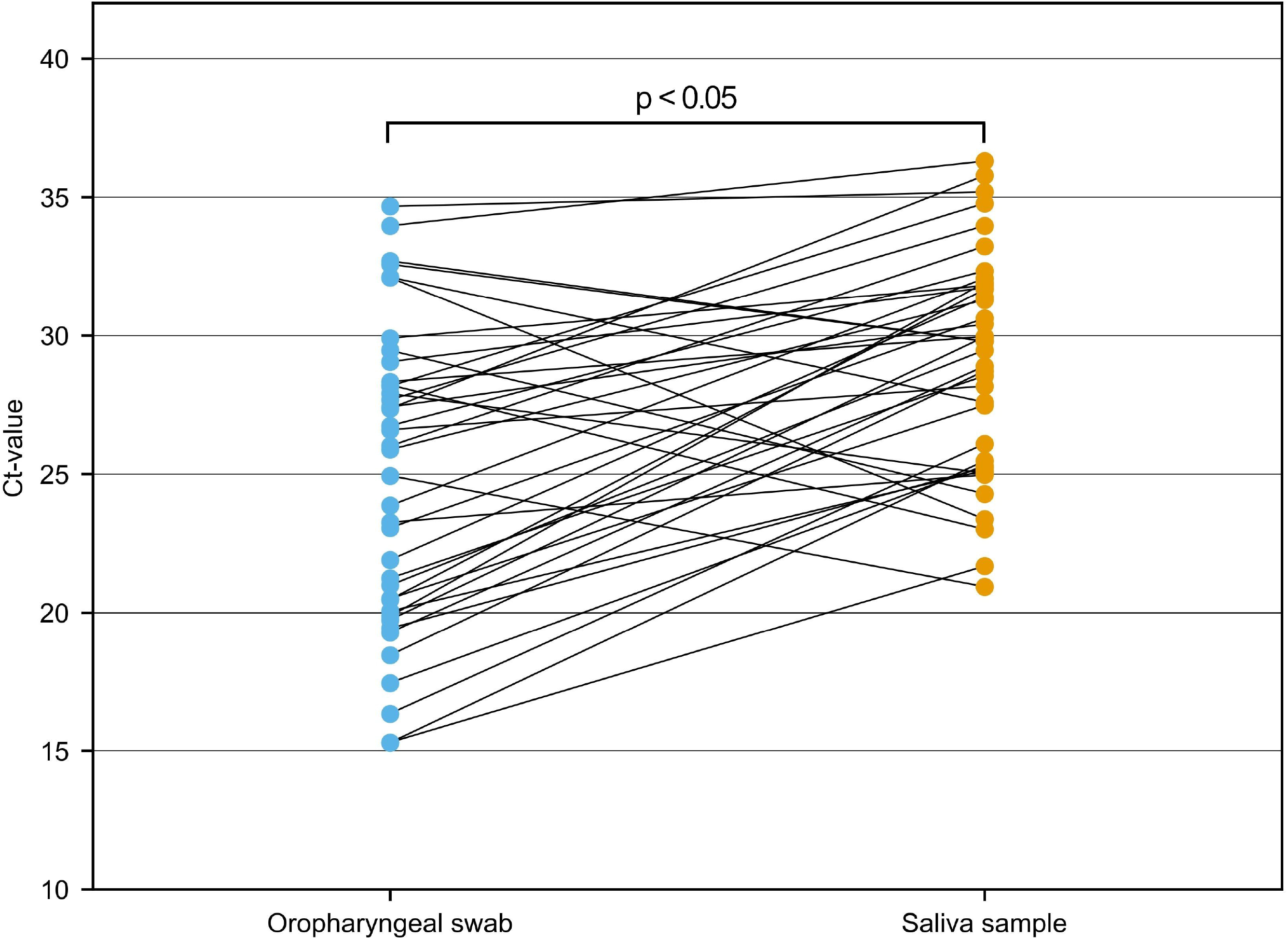
Comparison of Cycle threshold (Ct) values of SARS-CoV-2 rRT-PCR corresponding gene loci from 39 Patient-matched saliva and oropharyngeal swab samples (SC: 2, PC: 37); p-value was calcu-lated by Wilcoxon matched-pairs signed rank test.

**Figure 3.**
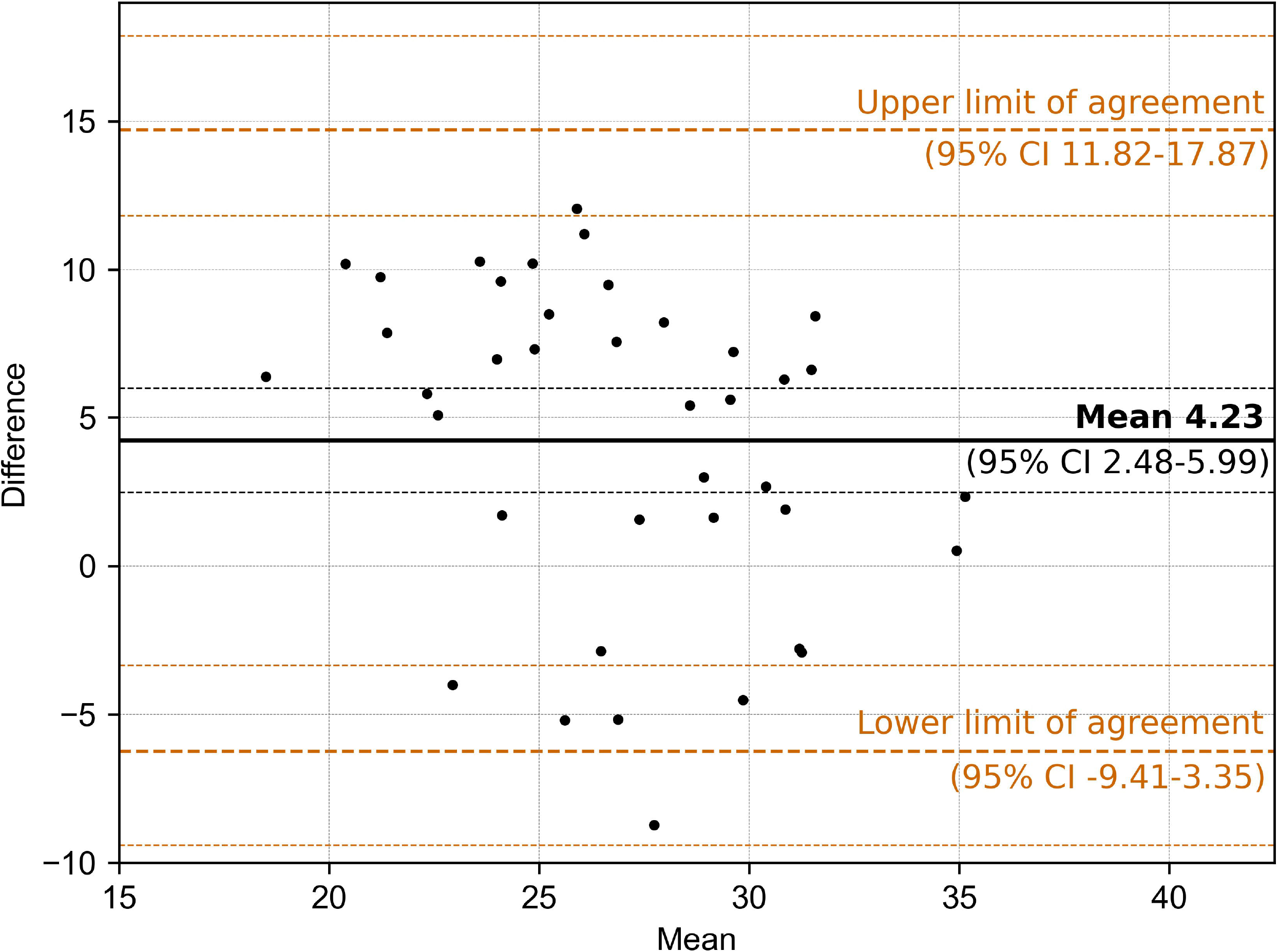
Bland-Altman graph displaying means and mean differences of Cycle threshold (Ct) values be-tween 39 saliva and oropharyngeal swab sample pairs (SC: 2, PC: 37) including upper and lower limits of agreement.

## Discussion

To our knowledge, this is the first large-scale feasibility study introducing the Salivette® system in combination with rRT-PCR for SARS-CoV-2 testing for children (aged 3 years and above) and staff in educational settings. So far only few studies reported the use of this testing algorithm in adults [19,24,25]. The Salivette® system is an easy-to-use, safe and feasible collection method licensed for supervised (professional use) saliva sampling in children aged 3 years and above. Its use for home-sampling in 201 adults over a 2-week period has been evaluated, comparing rRT-PCR results from saliva and oropharyngeal swabs [19]. Another recent study assessed Salivette® as a standardized saliva collection device and compared SARS-CoV-2 positivity with paired nasopharyngeal swabs and saliva specimens in about 300 adults. The authors conclude that, when using nasopharyngeal swabs as reference, Salivette® samples showed a sensitivity and specificity of 82.9% and 91.4%, respectively. But for samples containing less than 0.15 ml of saliva the Salivette® cotton roll was topped up with ultra-pure water and eluted again, introducing a relevant dilution effect prior to testing. This may explain higher sensitivity demon-strated in our study since we did not have to top up any Salivette® collected saliva sample and were thus always using neat saliva for SARS-CoV-2 testing.

We show that the mean difference in Ct-values between oropharyngeal swabs and saliva collected in the Salivette® system was significant (4.23). Still, overall test specificity of 100% and sensitivity of 94.9% in relation to standard swabs demonstrated in this study was excellent; of note, positive percent agreement between the two testing methods in indi-viduals with a high to moderate viral load (Ct-values < 33 from oropharyngeal swab samples) turned out to be 100%. This is of particular practical relevance since it proves that the Salivette® system is not inferior to oropharyngeal swab sampling in the most relevant group of individuals [26]. Some studies have demonstrated lower sensitivity and speci-ficity of saliva testing methods, but this is most likely due to inadequate pre-sampling conditions and sample volumes [19]. Most reports do not explicitly address important pre-analytic aspects [8,14,27]. They frequently remain unclear about the volume of saliva collected and whether saliva samples were processed as neat material or diluted (buffer or normal saline) in the laboratory before rRT-PCR testing. To address pre-analytic con-sistency, we measured the volumes of harvested saliva. The results showed a consistent amount of saliva in adults and children. It has previously been demonstrated that su-pervised sampling, like in our study, yields better results than self-collection or oropharyngeal washing [15]. Finally, saliva test results are likely to be influenced by prior fluid or food intake, smoking or other habits such as chewing gums. Melo Costa and colleagues recently assessed the concordance level between nasopharyngeal swab and Salivette® samples in 319 paired samples from adults. They found that routine mouth washes performed prior to obtaining saliva samples led to a substantial decrease in salivary viral loads thus negatively impacting on SARS-CoV-2 detection [22]. We ensured that saliva samples were not influenced by these factors. One may speculate that the best Salivette® sampling window would be when sampling is integrated as an early-morning, pre-breakfast and pre-toothbrushing routine procedure in the home setting. One limitation of our study is based on its design of two independent cohorts. While our study clearly demonstrates both feasibility and highly reliable test performance of the Salivette® system in the SC, the patient cohort for assessment of sensitivity was rather small. Comparison of Ct-values from concurrent sample pairs was only performed on 39 sample pairs. Since the SC proved to be a low incidence cohort with a low pre-test probability [4], we deliberately sought to establish the best cohort for assessment of sensitivity. Thus, our PC was characterized by a high pre-test probability. Hence it was only used to assess sensitivity and not designed to demonstrate specificity. In fact, specificity calculated from the PC would have shown a value of 80.0 %, but with an extremely wide 95% confidence interval (CI) ranging from 44.2% - 96.5% (Table 2b). In contrast, the 95% CI for specificity derived from the SC was 99.7% - 100% demonstrating a much more reliable result for specificity based on the SC. The four discordant results in the PC, a group of individuals all diagnosed with COVID-19 on a previous clinical rRT-PCR test result, are an important matter of debate. While the “saliva-negative / oropharyngeal swab-positive” pairs can easily be explained in view of the mean difference in Ct-values discussed above, we do not feel that the two “saliva positive / oropharyngeal swab-positive” pairs should be regarded as “false positive” since patients had a previously proven SARS-CoV-2 infection. In fact, we would interpret these positive results as “true positives” in view of consistent medical arguments. In a situation with SARS-CoV-2 infection subsiding in respective individuals (Ct-values 37.49 and 37.68), the Salivette® samples may have been superior to oropharyngeal swabs in detecting the virus due to an increasingly patchy distribution of the virus in the oropharynx. This issue of negative swab results and SARS-CoV-2 detection in saliva samples as a correlate for the persistence of the virus in the body after oropharyngeal swab conversion, has already been addressed and discussed by a number of groups [28,29]. Some authors even suggested to base evaluation of SARS-CoV-2 positivity on both oropharyngeal swab and subsequent saliva samples [29]. Our findings are further supported by a systematic review and meta-analysis comparing saliva and nasopharyngeal swabs for rRT-PCR testing for SARS-CoV-2 and demonstrating that both methods yield similar sensitivity and specificity across all 16 studies included in the analysis [13]. Other alternative non-oropharyngeal swab approaches have also been explored and may be practical for both adults and children. Whereas buccal swabs do not seem to be a reliable alternative option [30], Willeit and colleagues have reported promising results from gargling samples [31]. While this method may be feasible in adults and older children, it cannot be used in younger children. In addition, gargling involves external fluid or buffer whereas the Salivette® allows standardized collection of neat saliva as an undiluted clinical specimen. Furthermore, while Salivette®-based saliva collection, due to its closed system, is safe and not posing any risk for virus transmission to healthcare workers or friends and family nearby, gargling methods generate aerosols and are thus less suitable from an infection-control point of view. In view of recent evidence that SARS-CoV-2 also infects salivary glands and oral mucosa, saliva must be regarded as an optimal specimen of SARS-CoV-2 testing [32]. Furthermore, in situations where test capacities are limited the Salivette®-collected individual saliva samples can easily be pooled in the laboratory to assess 5 or more single samples in one rRT-PCR run [33]. Thus, we fully agree with Anne Wyllie’s group who recently concluded that a standardized, inexpensive, and broadly implementable saliva-based methods could make frequent, comfortable testing for SARS-CoV-2 a reality for communities globally [34].

## Conclusion

In view of the current pandemic situation with an increasingly rapid spread of coronavirus variants of concern and the B.1.617.2 (delta) variant in particular, a Salivette® based testing algorithm holds great potential for younger children, the largest yet unimmunized group of individuals, by ensuring safe operation of educational institutions.

## Supporting information

Age range of individual [years], sex and Ct-values of corresponding gene loci included in the head-to-head analysis (n=39)

## Data Availability

Data presented and discussed in the manuscript is available on request.

## Acknowledgements

Authors would like to thank Elisabeth Dick, Annalena Branz, Felix Flachenecker, Janina Ludwig, Adrian Rödig, Maria-Sophia Stadler, Jasmin Mahdawi and Johannes Nowak, Alexandra Köhler, Alexandra Schubö, Noah Lee and Maxim Ustinov for helping in the field and supporting editing processes. Authors would like to thank all participating institutions, their staff as well as all children and their parents for their valuable support. Authors would also like to thank the *Bayerisches Staatsministerium für Unterricht und Kultus*, the *Referat für Bildung und Sport der Landeshauptstadt München* for supporting our study. This study was approved by the ethics committee of the Ludwig-Maximilians University (LUM) under project number 20-484.

## Author’s contributions

MH, TS, SV and UvB developed the concept and designed the study, TS and UvB wrote study protocol. LK, TS and UvB obtained ethics approval. MH, SV, UE, LK, VG, SK, ARH, MMB and UvB contributed to data acquisition. UE, NA, AS, BL, MMB and VF supervised laboratory procedures. MH, SV, UE, LK, VG and UvB analyzed the data. MH, SV, LK, JH, TS and UvB interpreted the data. MH, SV and UvB wrote the manuscript. MH, SV, LK and UvB supervised development of illustrating figures and tables. All authors read and approved the manuscript and take full responsibility of its content.

