## Supplementary material for "Feasibility and diagnostic accuracy of saliva-based SARS-CoV-2 screening in educational settings and children aged < 12 years": Age range of individual [years], sex and Ct-values of corresponding gene loci included in the head-to-head analysis (n=39)

| **Supplementary table (S1):** Age range of individual [years], sex and Ct-values of corresponding gene loci included in the head-to-head analysis (n=39). | | | |
| --- | --- | --- | --- |
| **Age range of**  **individual** | **Sex (male, female)** | **Ct-value (SAL)** | **Ct-value (OPS)** |
| 21-25 | m | 20.94 | 24.94 |
| 46-50 | m | 21.68 | 15.3 |
| 66-70 | f | 23.01 | 28.21 |
| 21-25 | m | 23.37 | 32.10 |
| 46-50 | m | 24.29 | 29.46 |
| 56-60 | m | 24.97 | 23.26 |
| 21-25 | f | 25.03 | 27.9 |
| 26-30 | f | 25.13 | 20.05 |
| 21-25 | f | 25.23 | 19.43 |
| 46-50 | m | 25.48 | 15.29 |
| 51-55 | m | 26.09 | 16.34 |
| 6-10 | m | 27.48 | 20.51 |
| 71-75 | m | 27.59 | 32.11 |
| 31-35 | m | 28.17 | 26.6 |
| 21-25 | f | 28.54 | 21.24 |
| 31-35 | f | 28.72 | 18.45 |
| 46-50 | m | 28.89 | 19.28 |
| 31-35 | f | 29.47 | 20.99 |
| 61-65 | m | 29.79 | 32.58 |
| 71-75 | f | 29.79 | 32.7 |
| 56-60 | m | 29.95 | 19.74 |
| 11-15 | m | 29.96 | 28.33 |
| 31-35 | f | 30.41 | 27.42 |
| 36-40 | f | 30.62 | 23.06 |
| 21-25 | m | 31.29 | 25.88 |
| 31-35 | m | 31.39 | 21.9 |
| 66-70 | f | 31.67 | 20.47 |
| 86-90 | f | 31.73 | 29.05 |
| 66-70 | m | 31.81 | 29.9 |
| 31-35 | m | 31.92 | 19.87 |
| 76-80 | f | 32.07 | 23.86 |
| 46-50 | m | 32.34 | 26.74 |
| 21-25 | f | 33.97 | 27.68 |
| 46-50 | m | 34.78 | 28.17 |
| 1-5 | f | 35.19 | 34.67 |
| 76-80 | m | 36.3 | 33.97 |
| 16-20 | m | 25.30 | 17.45 |
| 11-25 | m | 33.23 | 26.01 |
| 6-10 | f | 35.78 | 27.36 |
| **Abbreviations:** SAL: Salivette® (Saliva); OPS: oropharyngeal swab. | | | |
